# Genome-wide association study of nociceptive musculoskeletal pain treatment response in UK Biobank

**DOI:** 10.1101/2022.01.04.22268719

**Authors:** Song Li, Geert Poelmans, Regina L.M. van Boekel, Marieke J.H. Coenen

## Abstract

Drug treatment for nociceptive musculoskeletal pain (NMP) follows a three-step analgesic ladder proposed by the World Health Organization (WHO), starting from non-steroidal anti-inflammatory drugs (NSAIDs), followed by weak or strong opioids until the pain is under control. However, effective pain treatment is challenged by inter-individual differences, and unsatisfied pain treatment response (PTR) rates ranging from 34 to 79% in those suffering from NMP. To investigate the underlying genetic component of PTR, we performed a genome-wide association study (GWAS) in ∼ 23,000 participants with NMP from the UK Biobank. In our primary analysis, we compared NSAID vs. opioid users as a reflection of (non)response to NSAIDs, adjusting for age, sex, BMI, population substructure, and study-specific covariates. One genome-wide significant hit was identified in an intergenic region on chromosome 4, rs549224715 (P = 3.88×10^−8^), and seven signals pass the suggestively significant threshold (P < 1×10^−6^). All identified loci were in non-coding regions, but most variants showed potential regulatory functions. SNPs in LD (r^2^ > 0.6) with the lead SNPs passing the nominal significant threshold (P < 0.05) were mapped to 28 target genes in FUMA. Eight of these 28 genes are involved in processes linked to neuropathic pain and musculoskeletal development. Pathway and network analyses with Ingenuity resulted in the identification of immunity-related processes and a (putative) central role of EGFR. Genetic correlation analysis including 596 traits resulted in the identification of 67 nominally significant (P < 0.05) genetic correlations, and these traits were significantly enriched for chronic pain and socioeconomic status traits (P = 3.35 × 10^−12^). Additionally, we conducted a subtype GWAS for inflammatory NMP and a secondary GWAS for participants with NMP disease history, but no significant hits or overlap with the primary analysis were observed. Overall, we identified one genome-wide significant association in this first GWAS focusing on pain treatment using the analgesic ladder as phenotype. However, we realize that this study lacked power and should be viewed as a first step to elucidate the genetic background of NMP treatment.

## 1. Introduction

Chronic musculoskeletal pain is one of the most frequent causes of suffering and disability, e.g., chronic back pain affects about 20% of the population worldwide per year [1]. The nature of musculoskeletal pain can be nociceptive or neuropathic, for which the corresponding pain management differs. To facilitate standardized pain management, the WHO advises a three-step analgesic ladder for cancer-related pain [2], which now also works as a framework for stepwise medical management of most nociceptive pain. The first treatment step is non-opioid analgesics, such as acetaminophen (paracetamol) or non-steroidal anti-inflammatory drugs (NSAIDs); the second step is weak opioids for mild to moderate pain, such as codeine or tramadol; the third step is strong opioids for moderate to severe pain, such as morphine or oxycodone.

Unfortunately, effective pain treatment is challenged by inter-individual differences, with unsatisfied pain treatment response (PTR) rates ranging from 34 to 79% [3]. The factors contributing to PTR differences are multifactorial, including demographic and anthropometric characteristics (age, sex, weight, race, socioeconomic status) [4-6], lifestyle (smoking and alcohol intake) [7], comorbidities (psychological status, liver and kidney diseases) [6,8], and genetic factors [9,10]. However, none of the above-described factors predict PTR sufficiently to be of value to optimize pain treatment in a clinical setting.

The genetic background of PTR has mainly been investigated using a candidate gene approach. These studies focused on genes coding for drug-metabolizing enzymes, opioid receptors, and genes implicated in pain and pain sensitivity. Evidence shows that some drug-metabolizing genes are associated with response to specific drugs, e.g., *CYP2D6* and codeine treatment outcome [11,12]. In addition, genes implicated in pain could also possibly contribute to PTR by interfering with pain development or the underlying disease. The most investigated genes in this category code for ion channels, cytokines, opioid receptors, and drug transporters [13]. Furthermore, genes involved in pain sensitivity may play a role in PTR because studies show that greater pain sensitivity is associated with increased opioid use [14] and poorer chronic pain treatment outcomes [15,16].

Except for a number of recent, successful large-scale genome-wide association studies (GWASs) of chronic pain phenotypes [17-21] and pain sensitivity [22], the number of genetic studies focusing on PTR is still limited. Candidate gene studies are limited in the number of genetic variants and sample size and report contradictory results [23,24]. Several studies investigating PTR focus on opioid receptor genes. Genetic variants in these genes are thought to influence the opioid response by altering µ-opioid receptor binding affinity of exogenous opioid ligands, such as morphine [25-27]. The most investigated genetic variant is the 118A to G basepair change in the OPRM1 gene. The G allele was associated with higher opioid dosing [28,29]. However, other studies showed that this allele was protective against pain [30,31]. Therefore, definitive conclusions on these genetic associations cannot be drawn yet. Moreover, the most frequently used phenotype in GWASs investigating PTR is acute pain (e.g., analgesic requirement or pain relief score after surgery [32-35]) but not long-term pain treatment outcomes.

This study sought to identify genetic variants associated with (long-term) PTR in people with NMP from the UK Biobank. A GWAS was performed including subjects treated according to the WHO pain treatment ladders, and comparisons were made between NSAID and opioid users as a reflection of PTR to NSAID.

## 2. Method

As our primary analysis, we conducted a GWAS of a binary phenotype comparing NSAID users and opioid users, using data from the UK Biobank.

### 2.1 Study population

The UK Biobank is a general population cohort with over 0.5 million participants aged 40–69 recruited across the United Kingdom (UK) [36]. Recently released primary care (general practitioners’, GPs’) data provides longitudinal structured diagnosis and prescription data, which was used for phenotype definition. Participants passing quality control (details below) were included for analysis. UK Biobank obtained informed consent from all participants.

### 2.2 Phenotype definition

To define the PTR phenotype, we first extracted all nociceptive musculoskeletal pain (NMP) treatments and diagnoses from the GP data. NMP diagnosis was primarily selected from the chapter on musculoskeletal and connective tissue diseases and relevant symptoms or signs from other chapters in the Read codes (versions 2 and 3). See Supplementary data 1 for the diagnosis codes included in this study. Secondly, pain prescriptions (NSAID and opioid) were extracted from the GP data using the British national formulary (BNF), dictionary of medicines and devices (dmd), and Read code (version 2) for data extraction. An overview of the extracted medication codes is provided in Supplementary data 2. Only participants with an NMP diagnosis record and a pain prescription record occurring on the same date were included for analysis to ensure that we would only include pain treatment for NMP.

Based on the information of NMP and pain prescriptions from the UK biobank, two PTR phenotypes were defined: an ordinal and a binary phenotype. A dichotomous score was used for the binary (case/control) PTR phenotype: NSAID users were defined as controls and opioid users as cases. Two additional quality control (QC) steps were applied. First, participants with only one treatment event were removed to safeguard the inclusion of only participants with relatively long-term treatment. Second, a chronological check was applied for the first prescription of each ladder to ensure that the treatment ladder was correctly followed, i.e., initial NSAID use was followed by weak opioids and then strong opioids. Participants that were not treated according to this order were removed. For the ‘ordinal phenotype’, an ordinal score of ‘1’, ‘2’, or ‘3’ was assigned to NSAID users (persons only using NSAIDs), weak opioid users (persons using NSAIDs and weak opioids), and strong opioid users (persons using NSAID, weak opioid, and strong opioids), respectively. Also for the ordinal analysis, patients with one treatment event and not following chronological treatment were removed.

### 2.3 Genotyping and Quality Control

Genotyping procedures have been described in detail elsewhere [37]. Routine QC steps for genetic markers on autosomes included removal of single nucleotide polymorphisms (SNPs) with (1) an imputation quality score less than 0.8, (2) a minor allele frequency (MAF) less than 0.005, (3) a Hardy-Weinberg equilibrium (HWE) test P-value less than 1 × 10^−6^, and (4) a genotyping call rate less than 0.95.

QC steps for genetic markers on the X chromosome’s pseudo autosome region (PAR) were the same as autosomes. For non-PAR of the X chromosome, additional QC steps included setting heterozygous haploid genotypes as missing for males, excluding multi-allelic SNPs, excluding variants with significantly different MAFs between males and females in the NSAID user group (P < 0.05/#SNPs), and variants that violated HWE were examined in the NSAID user group using only females.

Routine QC steps for the samples include removal of participants with (1) inconsistent self-reported and genetically determined sex, (2) missing individual genetic data with a frequency of more than 0.1, (3) putative sex-chromosome aneuploidy. Participants were also excluded from the analysis if they were considered outliers due to missing heterozygosity, not white British ancestry based on the genotype, and had missing covariate data.

### 2.4 Genome-wide association study

Based on the PTR phenotype definition, a different analysis strategy was used. For the primary analysis, we used the binary PTR phenotype (see **Fig. 1** for an analysis overview), and a GWAS was conducted using a linear mixed model in GCTA [38] for markers on the autosomeal chromosomes, adjusting for age, sex, BMI, depression history, smoking status, drinking frequency, assessment center, genotyping array, and the first ten principal components (PCs). The following variables from the UK Biobank data set were used for the covariate definition: (1) depression history, which was defined as “YES” if depression records were found in self-reported, inpatient hospital or GP data, and (2) drinking frequency, which was derived from data field 1558: “Daily or almost daily” or “Three or four times a week” was defined as high drinking frequency, other values except for “Prefer not to answer” were defined as low drinking frequency. Differences in categorical covariates were tested by a χ^2^ test. Differences in continuous covariates (age and BMI) were compared by t-test or ANOVA for the binary and ordinal phenotypes, respectively, in SAS version 9.4 (SAS Institute Inc., Cary, NC). To examine the nature of pain between groups, all NMP diagnosis codes were grouped into one of the following categories: inflammatory, mechanical, and mechanism not specified. The percentage of subjects in each diagnosis category was compared between groups by a χ^2^ test. For the ordinal PTR phenotype, an ordinal regression analysis was conducted in OrdinalGWAS [39], using the same covariates as for the binary analysis.

**Figure 1.**
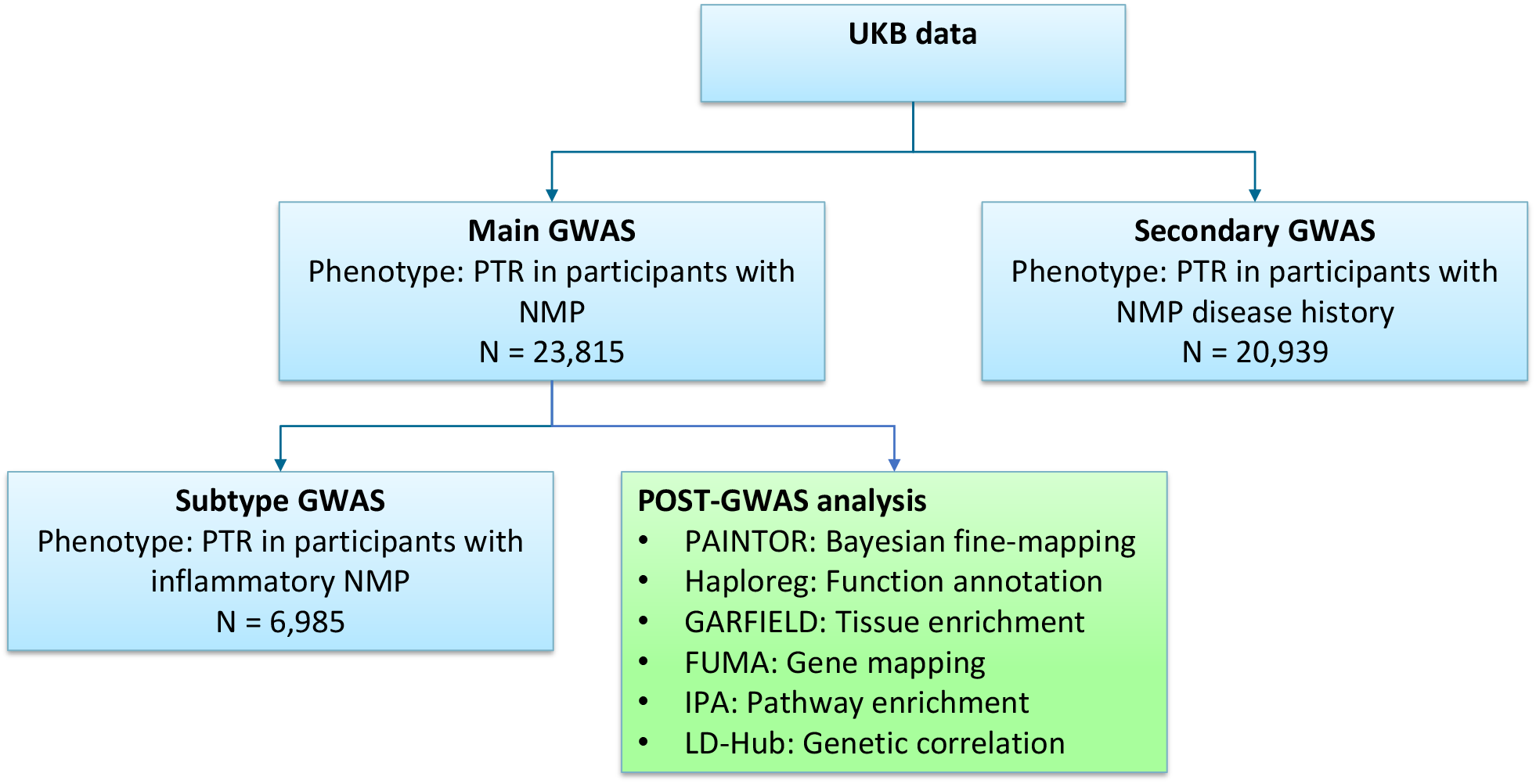
Overview of the study: Genome-wide association study for pain treatment response (PTR) in UK Biobank (UKB) participants with nociceptive musculoskeletal pain (NMP). Primary GWAS was conducted using the binary PTR phenotype: NSAID users vs. opioid users. Post-GWAS analyses were carried out for this primary analysis. Subtype GWAS was conducted including only participants with inflammatory NMP. Secondary GWAS was conducted including participants with pain prescription records and at least one treatment event for NMP, without limitations on treatment purposes. The blue boxes refer to the GWAS comparisons, and the green box indicates the post-GWAS methods

Association analysis of X chromosome markers was only examined for the binary PTR phenotype. Markers on the PAR region were tested using the same method as for the autosomal chromosomes, and a sex-stratified analysis was used for association analysis of the non-PAR region in XWAS [40].

A p-value less than 5 × 10^−8^ was considered genome-wide significant, and P-values between 1 × 10^−6^ and 5 × 10^−8^ were defined as suggestively significant. Narrow-sense heritability was only calculated using data from the primary analysis (binary phenotype) using the GREML method in GCTA [38,41]. FUMA was used to define the lead SNPs (SNPs with the smallest P-value in each locus) and independent, significant signals (SNPs remaining significant after conditioning on the lead SNPs in each locus) [42].

The power for the primary GWAS was calculated using the CaTS power calculator (http://csg.sph.umich.edu/abecasis/cats), assuming an additive model with the following input parameters: a significance level of 5 × 10^−8^, the prevalence of phenotype (opioid use) of 50%, and a relative genotypic risk of 1.135, based on 11,089 cases and 12,726 controls.

### 2.5 Functional annotation

Only the results of the primary analysis were carried forward for the following post-GWAS analysis.

#### 2.5.1 Bayesian fine-mapping of lead loci

Lead SNPs from the primary analysis were analyzed using Bayesian fine-mapping in PAINTOR [43] to identify the most likely causal SNP in each locus. PAINTOR calculates the posterior probability (PP) of causality for SNPs in each genomic region by leveraging the strength of association (Z score) and the LD pattern. The 1000 Genomes (Phase 3) were used for LD matrix calculation. The calculated PP for each SNP was sorted from high to low, and variants together reaching a PP of 0.95 were used to define 95% credible sets.

#### 2.5.2 Functional annotation of SNPs present in the 95% credible sets

SNPs in the 95% credible sets were annotated for regulatory functions in HaploReg v4.1 [44]. The analyzed regulatory functions were (1) the presence of exonic, nonsynonymous variants in high LD (r^2^ ≥ 0.8), (2) overlap with epigenetic histone marks of active enhancers (H3K4me1 and H3K27ac) and active promoters (H3K4me3 and H3K9ac), and (3) the sensitivity to DNase. As histone marker overlap is tissue-specific, relevant cell lines were selected from the complete data set (see Supplementary Table 1). Besides regulatory functions, potential pleiotropy effects of the variants were investigated. For this, it was investigated whether the SNPs from the 95% credible sets were found associated with other phenotypes by checking previously reported association with other phenotypes in Haploreg. For SNPs not available in Haploreg, proxy SNPs were obtained by LD proxy (https://ldlink.nci.nih.gov/). For loci containing more than ten likely causal variants, only the lead SNP and SNPs with the maximum posterior probability (PPmax) of the SNPs in one locus were annotated.

#### 2.5.3 Epigenetic marks enrichment analyses

GARFIELD was used to investigate tissue-specific regulatory and functional enrichment of the GWAS signals [45]. GARFIELD firstly pruned SNPs in the summary statistics results to obtain independent GWAS SNPs (r^2^ < 0.1). Then, the independent SNPs were annotated for functional overlap with a panel that includes 1005 annotations (genetic annotations, chromatin states, histone modifications, DNase I hypersensitive sites, and transcription factor binding sites in different cell lines) derived from the ENCODE, GENCODE, and Roadmap Epigenomics projects [46-48]. The interpretation of the functional consequences was conducted in each cell line and primary tissue. Next, the significance of the functional enrichment was tested at GWAS significance cutoffs (1 × 10^−5^) under a generalized linear model, adjusting for significant sources of confounding factors, including minor allele frequency, distance to nearest transcription start site, and the number of LD proxies (r^2^ > 0.8). Multiple testing correction was applied for an enrichment P-value threshold (0.05/Meff) based on the effective number of annotations (Meff = 525.95).

#### 2.5.4 Gene mapping

In FUMA, three strategies were adopted to map suggestively significant (P-value < 1 × 10^−6^) GWAS SNPs and SNPs in LD (LD> 0.6) with them to genes: positional mapping, expression quantitative trait loci (eQTL) mapping, and chromatin interaction mapping. For the positional mapping, it was investigated if the SNPs mapped to known protein-coding genes based on physical distance (within a 10 kb window). For eQTL mapping, it was investigated if SNPs mapped to genes up to 1 Mb away based on cis-eQTLs. As gene expression is tissue-specific, we selected the following tissues for mapping: brain, muscle, kidney, liver, nerve, skin, and fibroblast. Significant eQTLs were defined as eQTLs with a false discovery rate (FDR) < 0.05. Finally, chromatin interactions were assessed. Chromatin interaction can occur in two genomic regions that are spatially close when DNA folds together, even if the genomic regions are in long-range physical distance. Genes in regions of chromatin interaction containing candidate SNPs were assessed in the same tissues as the eQTL mapping. An FDR < 1 × 10^−6^ was defined as a significant interaction.

### 2.6 Pathway enrichment analysis

To investigate if the genes identified in the GWAS could be linked to biological pathways and networks involved in PTR, a gene-based functional pathways enrichment was performed using Ingenuity Pathway Analysis software (IPA®, QIAGEN Redwood City) [49]. IPA is based on prior knowledge of direct and indirect gene relationships from experimentally observed data in mammals and all cell types. A gene-based P-value was computed twice using the gene analysis function in MAGMA v1.08 [50]. Firstly, a gene-based P-value was calculated using nominally significant SNPs (P-value < 0.05) in the protein-coding region of genes without flanking regions. Secondly, the same analysis was performed including only nominally significant SNPs in the protein-coding regions of gene, as well as in 100 kilobase (kb) pair upstream and downstream flanking regions of genes, to take cis-eQTL effects into account [51-54]. Both analyses were combined, and the smallest P-value of each gene was selected for pathway and network analyses with IPA (see gene list in Supplementary Table 2). Pathways with an FDR < 0.05 were considered as statistically significantly enriched. To illustrate the core molecules in the networks, a radial plot was generated of the merged top five networks.

### 2.7 Genetic correlation analysis

Genetic correlations between PTR and other complex traits were investigated by linkage disequilibrium score regression through LD Hub v1.9.3 [55]. The tested traits were selected from LD hub, and the following categories were selected: education, anthropometric, sleeping, psychiatric, personality, cognitive, autoimmune, smoking behavior, kidney, neurological, and UKBB phenotypes. Correlations with P-values less than 8.4 × 10^−5^ (0.05/596) were considered significant. Since the top nominally significant correlations were overrepresented by pain and socioeconomic status phenotypes, the percentage of nominally significant (P < 0.05) correlations in these two categories was compared with all the other categories by a χ^2^ test. The socioeconomic status phenotypes consisted of the pain category (category 100048), medication for pain relief, constipation, heartburn (data field 6154), qualifications (data-field 6138), and employment (category 100064) in the UKB and all education phenotypes in LD hub [56-59].

### 2.8 Subtype GWAS and Secondary GWAS

As inflammatory pain is an important subtype of pain, a subtype GWAS was carried out for this phenotype specifically. Only inflammatory NMP diagnosis codes were used for participant selection. Some participants with inflammatory NMP received pain treatment for other types of pain (e.g., mechanical NMP or not specified) over time. These participants were excluded from the analysis.

Moreover, as diagnostic codes were often not repeatedly recorded [60] or reported as repeat prescriptions, a secondary GWAS was performed for pain medication users with less strict diagnosis criteria than the primary analysis. To ensure a relatively homogenous population, we included participants with at least one NMP treatment event but without any other limitations on their pain treatment purposes. To focus on long-term treatment outcomes and remove outliers, participants were removed if they only had one prescription record or prescription record numbers on a log scale outside the 1.5 inter-quantile range. In addition, participants already included in the primary GWAS analysis were not included in this analysis.

Both the subtype and secondary GWASs were performed using the binary PTR phenotype and following the same procedure as the primary analysis, but only autosomal markers were examined. To investigate whether the identified loci in these two analyses overlapped with the primary analysis, lead SNPs with a suggestive threshold (P < 1 × 10^−6^) were compared with the primary GWAS signals for LD correlation (r^2^ > 0.6) in LDpair (https://ldlink.nci.nih.gov).

## 3. Results

### 3.1 Primary GWAS

After quality control, we identified 12,592 NSAID users, 10,341 weak opioid users, and 365 strong opioid users in the UK Biobank dataset. Considering the small sample size of the strong opioid group (n = 365) and assuming mechanistic similarities between weak and strong opioids, we decided to combine these two opioid groups, leading to 12,726 NSAID users and 11,089 opioid users. The sample size of both groups was increased after combing the weak and strong opioid users because it was possible to include related individuals in a linear mixed model, and participants taking strong opioids directly after NSAIDs were also included in the opioid users group.

A case-control approach was used for the primary GWAS analysis: NSAID users (controls) versus opioid users (cases). **Table 1** summarizes the demographics of the cases and controls, and all tested covariates were found to be significantly different (P < 0.0001) between cases and controls. As the percentage of patients with inflammatory pain was significantly different between the two groups, we decided to run a subtype GWAS including only this subgroup with inflammatory pain (see below).

There were 9,435,994 SNPs available for GWAS analysis after quality control. Initially, we intended to fit a linear mixed model for the binary PTR phenotype. However, as the number of closely related participants was low, the linear function in GCTA was automatically adopted. The genomic control value (lambda) was 1.008. One intergenic locus located at chromosome 4 reached genome-wide significance, in which the most significant SNP was rs549224715 (P = 3.92 × 10^−8^) **(Fig. 2, Table 2)**. Seven SNPs in seven other loci surpassed the suggestive P-value threshold (P < 1 × 10^−6^), and no other independent SNPs (SNPs remaining significant after conditioning on lead SNPs in the locus) were identified in each locus. The SNP heritability was 0.16 (P-value = 0.16). In addition, an ordinal GWAS was conducted, and the results were consistent with the results of the primary GWAS (Supplementary Fig. 1A, Supplementary Fig. 2A).

**Figure 2.**
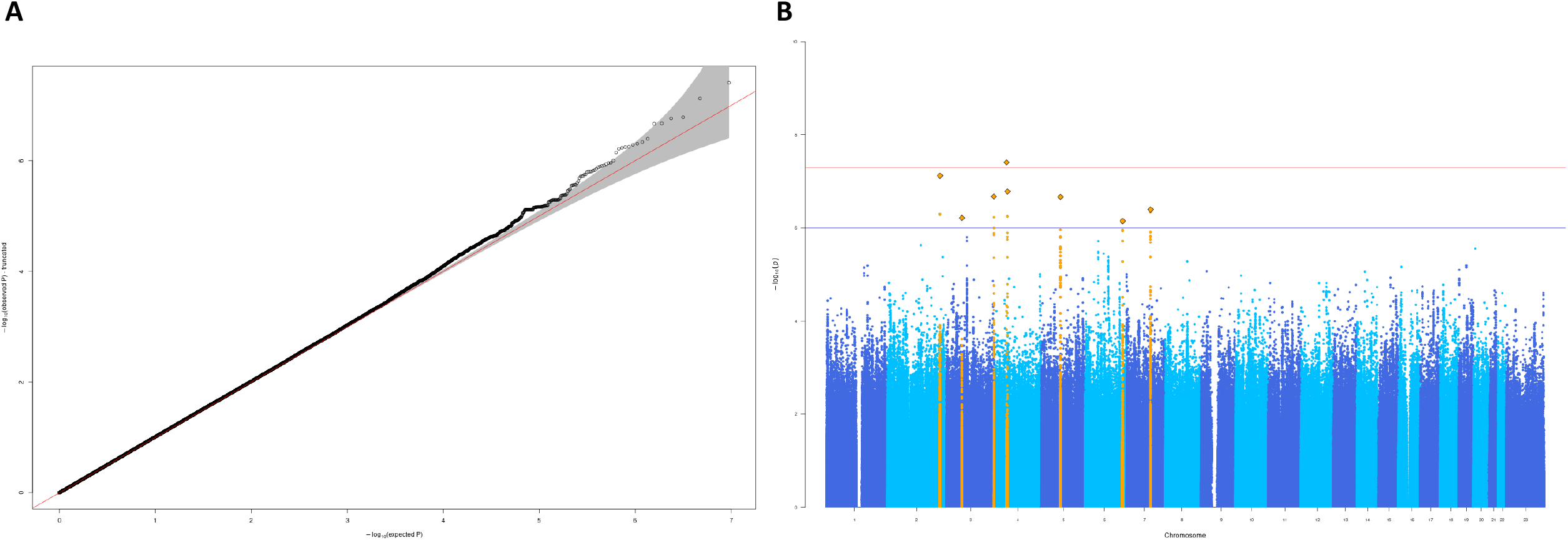
Q-Q plot and Manhattan plot of primary analysis for PTR. (A) Q-Q plot of the primary GWAS. The red line indicates the distribution under the null hypothesis, and the shaded area indicates the 95% confidence band. (B) Manhattan plot of the primary GWAS results. The red line corresponds to the genome-wide significance threshold 5 × 10^−8^, whereas the blue indicates the suggestive threshold to 1 × 10^−6^. Lead variants are highlighted as orange diamonds. Variants in one locus (within 400 Kb) are highlighted in orange.

### 3.2 Functional annotation

#### 3.2.1 Statistical fine-mapping of loci and functional annotation of SNPs

As GWAS signals can be caused by SNPs in linkage disequilibrium with the likely causal SNPs, we calculated the posterior probability for variants in each genomic locus and created 95% credible sets (see methods). In five out of eight loci, the lead SNPs in the locus had the maximum PP (PP_max_) (Supplementary Fig. 3).

Since all variants in the 95% credible sets were in non-coding regions, the regulatory effects of these variants were investigated by examining overlap with epigenomic markers of active enhancers or promoters in Haploreg. The results suggested that most genetic variants were potentially involved in transcriptional regulatory modulation (Supplementary Fig. 3).

We assessed whether the SNPs in the 95% credible sets were previously reported to be associated with other traits. However, no pleiotropic effects were identified.

#### 3.2.2 Regulatory element enrichment

To estimate functional element enrichment in specific tissues, we used GARFIELD to quantify fold enrichment (FE) for the genetic variants with a P-value < 1 × 10^−5^ in DNase I hypersensitivity site (DHS) hotspots in 424 cell types from different tissues. The significance threshold for enrichment was set at P < 9.5 ×10^−5^ (Bonferroni correction for the effective number of independent annotations (525.95)). We did not identify any significant enrichment (Supplementary Fig. 4). The highest enrichment was found for urothelium (FE = 3.98, P = 0.005), fetal muscle (FE =3.05, P = 0.026), and skin tissues (FE =2.72, P = 0.045).

#### 3.2.3 Gene mapping

After mapping GWAS candidate SNPs (SNPs that are in LD (r2 > 0.6) of any independent significant SNPs) to genes, a total of 28 unique mapped genes were identified (**Table 3**). Five genes were mapped by genomic location, nine genes were identified by cis-eQTL mapping, 18 genes were annotated as SNPs in regions where 3D chromatin interactions occurred, and four genes were identified by at least two mapping strategies.

### 3.3 Pathway enrichment

Pathway enrichment analysis in IPA prioritized 15 significant pathways with an FDR < 0.05, in which the top prioritized pathways were mainly implicated in the immunological response. (Supplementary Table 3).

The network analysis yielded a total of 25 prioritized networks. The top network contained 33 genes with the EGFR protein in the center. EGFR remained in the center after merging the five networks with the lowest P-value (Supplementary Table 4, Supplementary Fig. 5).

### 3.4 Genetic correlation with other traits

The genetic correlation analysis did not yield significant correlations (Bonferroni corrected P-value < 8.39 × 10^−5^). The top correlated trait was overall health rating (rg = 0.5316, P = 0.0087), followed by years of schooling [57] (rg = -0.5431, P = 0.0102) (Supplementary Table 5). However, among the nominally significant correlations (P < 0.05) we found an overrepresentation of pain and socioeconomic status traits compared to the other traits (43.48% vs. 8.55%, P = 3.35 × 10^−12^, Supplementary Table 6).

### 3.5 Subtype and secondary GWAS

As inflammatory (i.e., inflammation-related) pain is an important subtype of nociceptive pain, we performed a stratified GWAS in this more homogenous population. In this subtype GWAS, a total of three suggestively significant loci were identified, but none of them were in LD with the SNPs identified in the primary GWAS (Supplementary Fig. 1B, Supplementary Fig. 2B, Supplementary Table 7).

A secondary GWAS was performed in participants with pain prescriptions records but with less strict criteria for diagnosis. In this analysis, three signals passed the suggestive threshold for significance, but these signals did not overlap with the primary GWAS results either (Supplementary Fig. 1C, Supplementary Fig. 2C, Supplementary Table 8).

## 4. Discussion

To our knowledge, this is the first GWAS reporting on long-term pain treatment outcomes. We identified one genome-wide significant hit and seven loci with suggestive significance. Although pain or PTR is characterized by sex differences, i.e., females are more vulnerable to pain and opioid use [61], no significant signals were found on the X chromosome. The subtype and secondary GWAS analysis did not replicate the initial findings and did not yield additional loci associated with PTR.

This study is the first to report the narrow-sense heritability of the pain treatment response to NSAIDs. Although the heritability analysis result is not significant, it indicates a moderate heritability of PTR. Since there are no previous results of PTR heritability, we examined whether the heritability in our study is comparable to the heritability of response to opioid analgesics and chronic pain. The heritability in our study is in agreement with the heritability of opioid response (60% in cold pressor induced pain and 12% in heat pressor) in a twin study [62] and that of chronic pain (0.08 to 0.31) [8,17-21]. In addition, the possible reason for the low heritability calculated in this study could be that we were only able to calculate the narrow-sense heritability that captures the additive genetic components of common variants without the contributions of non-additive effects, rare variants, and structural variants. The other reason could be that PTR is a highly complex phenotype with other contributing factors such as employment status and psychological factors [63].

One genome-wide significant SNP was identified, rs549224715, but the link of this SNP with PTR remains unclear. The nearest gene is *CWH43*, which is 24 Kb away from this SNP. This gene is associated with Seckel Syndrome, characterized by growth delays prior to birth. Another gene, *TXK*, was mapped by eQTL to this SNP, and encodes a member of the non-receptor tyrosine kinases of the Tec family that plays a role in the regulation of the adaptive immune response [64]. The functional link between this SNP and PTR is worth further validation and investigation.

Most of the variants identified within the composition of the 95% credible sets showed potential transcription regulatory functions. This aligns with research indicating that epigenetic changes are involved in chronic pain [65,66] and pain treatment [67]. Some preliminary published results indicate that epigenetic restructuring can happen in response to opioid analgesics use. For instance, hypermethylation in both the promoter region of a candidate gene (*OPRM1)* and global DNA methylation were observed after opioid use [68,69].

In total, we pinpointed 28 genes that could be linked to the identified SNPs based on physical, eQTL, and chromatin interaction mapping. Four of the identified genes are of interest as these are involved in neuropathic pain: *NPTX2, THBS4, HOMER1*, and *IPCEF1. NPTX2* was identified by both eQTL mapping and gene-based analysis with the lowest P-value (2.71 × 10-5) (Supplementary Table 2). This gene encodes a member of the neuronal pentraxins family, which are involved in excitatory synapse formation. *NPTX2* is also thought to play a role in anxiety [70] and Alzheimer’s disease [71]. Recent evidence shows that *NPTX2* is downregulated in the brain in induced chronic neuropathic pain [72] and induced endometriosis [73] mouse models. The second gene implicated in neuropathic pain is *THBS4*, and this gene encodes a protein belonging to the thrombospondin protein family, involved in local nervous system signaling during development and adulthood [74]. Furthermore, THBS4 contributes to spinal sensitization and neuropathic pain states after peripheral nerve injury [75,76]. HOMER1 is a neuronal protein that regulates glutamatergic signaling in the brain [77] and interacts with Group I metabotropic glutamate receptors (mGluR1/5) in the postsynaptic density [78]. Glutamate receptor/Homer signaling is crucially involved in synaptic plasticity associated with chronic inflammatory pain [79,80] and neuropathic pain processing after nerve injury [81,82]. Lastly, IPCEF1 has been implicated in nerve injury-induced membrane receptor trafficking in dorsal root ganglions (DRG) in neuropathic pain conditions [83]. In addition to the genes linked to neuropathic pain, we identified four genes that could be linked to muscular or skeletal dystrophy; *CMYA5* [84,85], *SGCB* [86], *TMEM130* [87], and *FN1 [88,89]*. These genes are of interest as musculoskeletal dystrophy is characterized by pain.

In addition, no candidate genes were implicated in the metabolism and working mechanisms of pain medications. This could indicate that participants are more likely to use opioids because of pain or disease progression. However, our results do not necessarily exclude the role of genes involved in drug absorption, distribution, metabolism, and excretion (ADME) in PTR. One reason is that the subcategories in NSAIDs, such as non-selective NSAIDs and selective COX-2 inhibitors, were analyzed as a whole in this analysis, which may dilute their effect in the PTR difference. To overcome this, stratified analyses per drug should be performed. However, this was not possible as the groups would become too small to obtain sufficient power. That no variants involved in opioid processing were identified is in line with our expectation because the phenotype we used is a proxy for the treatment response to NSAIDs. The other possible reason is that rare variants in ADME genes can contribute significantly to treatment response differences [90]. Our study had 80% power to identify SNPs with a risk allele frequency of 5% and genotypic relative risk of 1.135, but we lack the power to detect variants with less than 5% frequency or smaller effect sizes. Therefore, it would be interesting to investigate the effect of rare variants of genes in a larger sample with a Next-Generation Sequencing-based method.

Although no correlations with PTR remained significant after Bonferroni correction, the enrichment of top correlations with nominal significance was perhaps expected. The overrepresentation of pain phenotypes is in line with literature indicating that opioid users tend to have more chronic and severe pain conditions [91]. The correlation with education and occupation also matched the reports that people carrying out strenuous occupations (jobs involving heavy manual or physical work) are more likely to report pain [18-20,92]. Our study indicates that people with strenuous occupations are more likely to have adverse treatment outcomes, i.e., to require analgesics at a higher step in the treatment ladder.

Identifying the relevant tissues is important for elucidating the etiology of pain and for further functional validations. Tissue enrichment using GWAS results may better reflect pain development under pathophysiology conditions than induced pain [93] in preclinical studies using animal models. Although our GWAS results are underpowered for tissue enrichment analysis, the top loci with suggestive threshold could still point to relevant tissues. The top enriched tissues include muscle tissue, which coincides with the identified genes involved in muscular dystrophy. These results probably stress the role of muscular tissue in pain progression as muscle pain is mediated by free nerve endings in muscle tissue [94]. However, these results are underpowered and warrant further validation.

The pathway enrichment and network analysis should be interpreted carefully as the input consisted of nominally significant genes from the underpowered primary GWAS analysis. Top prioritized pathways were mainly implicated in immunity-related processes. One of the pathways identified was retinoic acid-mediated apoptosis signaling. Although there seems to be a link between retinoic acid-mediated apoptosis signaling and pain, the results in the literature are not consistent. It has been reported that retinoic acid (RA) administration can reduce chemotherapy-induced experimental neuropathy [95] or inhibit prostaglandin synthesis in astrocytes [96], an important mediator of inflammation and pain signaling. In contrast, topical application of RA can induce retinoid-elicited irritation [97] by nociceptive pathways sensitization [98,99] or ionotropic receptor TRPV1 [100]. The network analysis emphasizes the role of EGFR (a member of the ErbB family of receptors) in PTR. Some links between EGFR and pain can be found in the literature. For instance, EGFR inhibition can block inflammatory chronic pain progression in preclinical studies [101] and relieve neuropathic pain in clinical settings [102-104], suggesting that the role of EGFR in PTR is worth further investigating.

Our subtype GWAS focusing on inflammatory pain did not identify genes linked to inflammation, nor did it strengthen the associations found in our primary GWAS. This could be explained by the loss of power due to the decreased sample size, and it could also indicate that inflammatory factors may not be a predictor for the severity of NMP [105]. The secondary analysis was aimed at validating the findings from the primary GWAS in a more heterogeneous group but unfortunately, we were unable to replicate the findings from the primary analysis.

By utilizing UK Biobank data, a large sample size with long-term pain treatment outcomes was available using a derived phenotype as a proxy for the pain treatment response to NSAIDs. However, it is difficult to validate this derived phenotype and assess pain chronicity because of a lack of appropriate diagnosis codes for pain in the current International Classification of Diseases (ICD) [83]. Therefore, we had to simplify our outcome definition of pain treatment. In clinical practice, pain treatment starts from paracetamol, then moves to NSAIDs, weak opioids, strong oral opioids, and finally strong intravenous opioids. Although applying this strict treatment ladder as phenotype can capture more phenotypic variance, only about 1000 participants started with paracetamol as their first treatment. Therefore, we had to combine paracetamol and other NSAIDs as the first treatment step. For the same reasons, oral and intravenous strong opioids were combined as the third step. In addition, our analysis assumes unidirectional analgesic ladders, whereas this can be bidirectional in clinical practice. Despite the limitations on phenotype definition, the group characteristics are similar to previous publications, with a roughly even share of NSAID users and opioid users in the population [106], and the reported risk factors for using opioids are also in line with previous literature [91,106,107]. Another point worth noting is that we did not control for adjuvant analgesics (or co-analgesics). Adjuvant analgesics, such as antidepressants, are used in each analgesic ladder as multimodal analgesia when patients demonstrate a poor response to analgesics. Although applying adjuvant analgesics can potentially influence pain treatment outcomes, we did not control for it in our analysis because of the limited clinical evidence for the use of most adjuvant analgesics [108] and the difficulties in distinguishing the primary indications of adjuvant analgesics in the data.

Unfortunately, replication of the results is difficult due to the limited number of publicly available large-scale data similar to UK Biobank and the lack of cohorts measuring long-term PTR. As pain treatment ladders can also be applied to other forms of chronic pain, we checked whether the data for pain medication users for orofacial and visceral pain in the UK Biobank cohort were sufficient to perform a validation analysis. However, only about 1,300 participants were found in total. Considering the sample size, we decided not to carry out this analysis. However, it is still worth exploring the genetic background of PTR in a large cohort with a specific PTR definition, such as the ongoing Pain Predict Genetics cohort in our center (NCT02383342).

In conclusion, we identified one locus achieving genome-wide significance for a derived pain treatment response phenotype. Some of the identified genes could be linked to neuropathic pain and musculoskeletal development. However, this study should be viewed as an initial stepping stone for future research.

## Data Availability

All data produced in the present study are available upon reasonable request to the authors.

## Acknowledge

The authors thank Ward De Witte for assistance with data analysis. This research has been conducted using the UK Biobank Resource under Application Number 52524. The authors are grateful to the UK Biobank participants for making such research possible.

This work is part of the research programme Computing Time National Computing Facilities Processing Round pilots 2018 with project number 17666, which is (partly) financed by the Dutch Research Council (NWO). This work was carried out on the Dutch national e-infrastructure with the support of SURF Cooperative.

S.L. was supported by China Scholarship Council (CSC) Grant number 201908130179.

Summary statistics of the primary analysis are available at DANS archive.

Gene mapping results are available at FUMA (https://fuma.ctglab.nl/browse/378).

## Table footnote

Table 1. Demographics of NSAID users (control) and opioid users (cases) in the UK Biobank. Categorical covariates are represented as frequency (percentage) and compared by the χ^2^ test. Continuous covariates are presented as mean (standard deviation) and compared by independent t-test. Depression history was defined as “YES” if depression records were found in the self-reported, inpatient hospital, or GP data. Drinking frequency was defined from data field 1558, “Daily or almost daily” or “Three or four times a week” defined as high drinking frequency, other values except for “Prefer not to answer” defined as low drinking frequency. * Percentage of subjects within a certain category of pain in each group. Note that one subject could have more than one type of diagnosis, so the sum of percentage is not equal to 1.

Table 2. Overview of the lead SNPs passing suggestive significance in the primary GWAS for pain treatment response. Bold font indicates the SNP that passed the genome-wide significant threshold (5 × 10^−8^). CHR:POS physical position of the SNP, A1 effect allele, AF1 effect allele frequency, BETA (SE) effect size of SNP and standard error (SE).

Table 3. SNPs mapped to candidate genes using FUMA. SNPs in LD (r^2^ > 0.6) with lead SNPs were mapped to genes. posMapSNPs, the number of SNPs mapped to this gene based on positional mapping; eqtlMapSNPs, the number of SNPs mapped to this gene based on eQTL mapping; eqtlMapminP, the minimum eQTL P-value of mapped SNPs; eqtlMapminQ, the minimum eQTL FDR of mapped SNPs; ciMap, “Yes” if the gene is mapped by chromatin interaction mapping; minGwasP, the minimum P-value of mapped SNPs.

