## supplementary figures for "Genome-wide association study of nociceptive musculoskeletal pain treatment response in UK Biobank"

### Slide 1
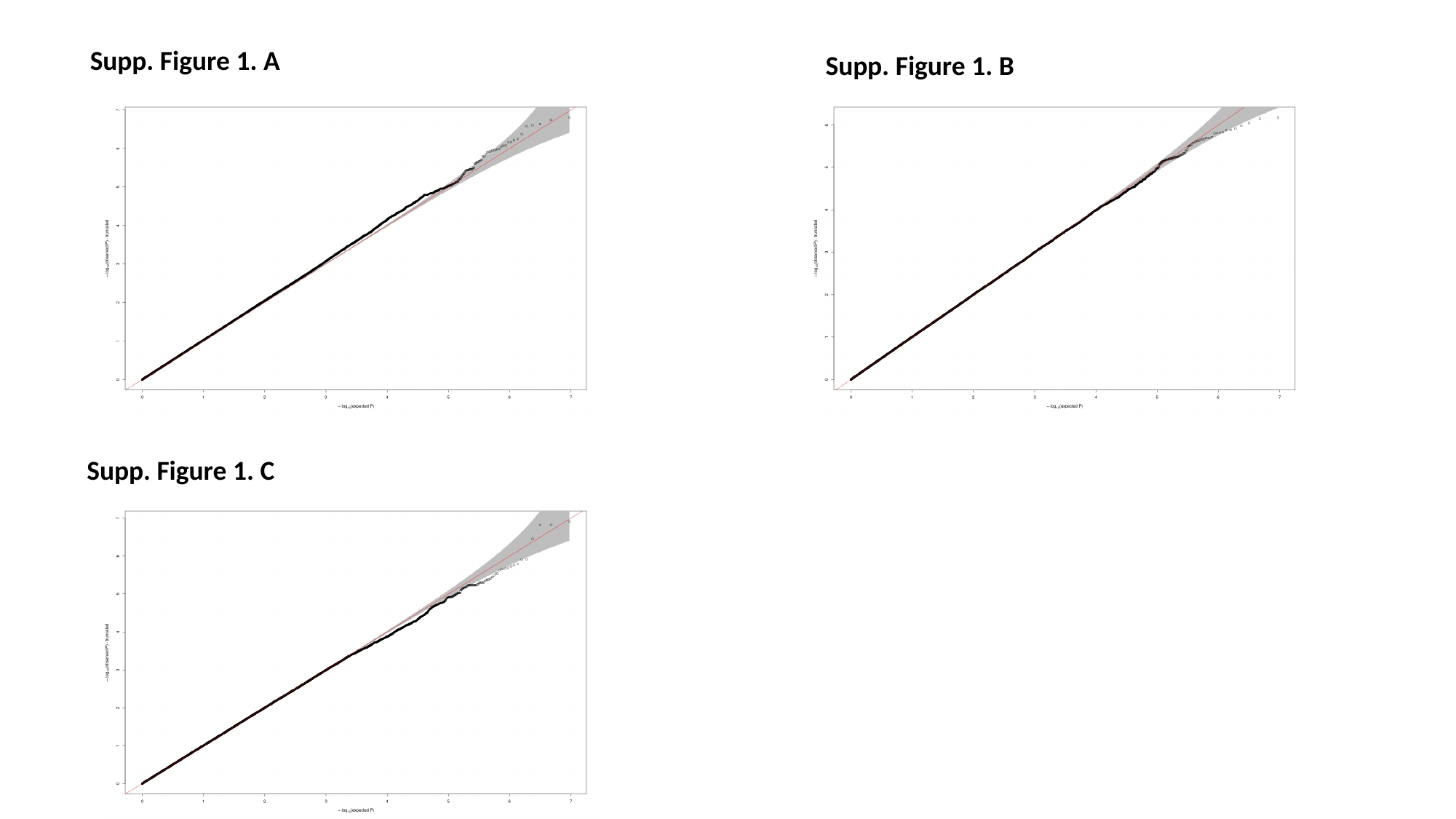

Supp. Figure 1. A
Supp. Figure 1. B
Supp. Figure 1. C

### Slide 2
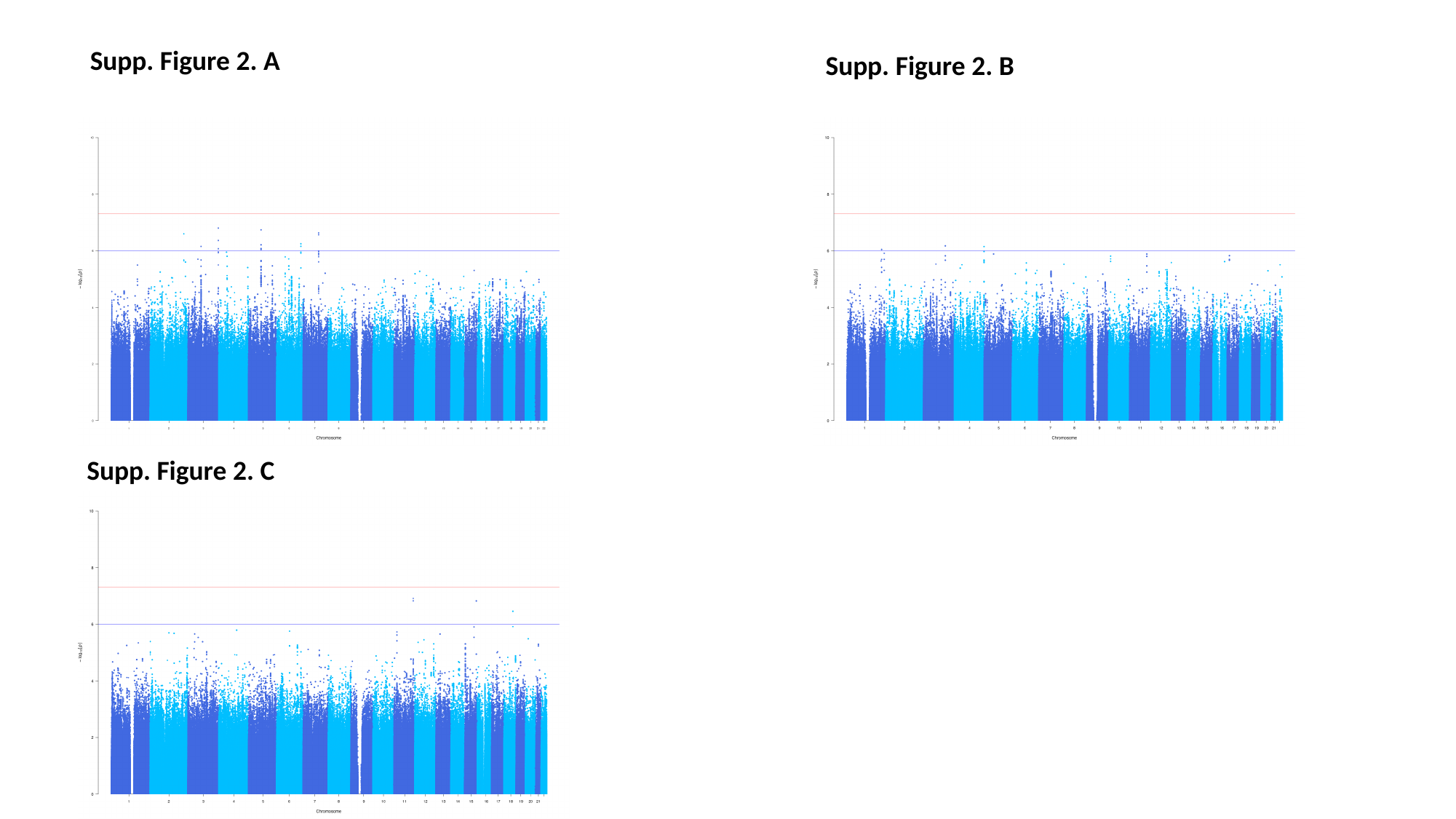

Supp. Figure 2. A
Supp. Figure 2. B
Supp. Figure 2. C

### Slide 3
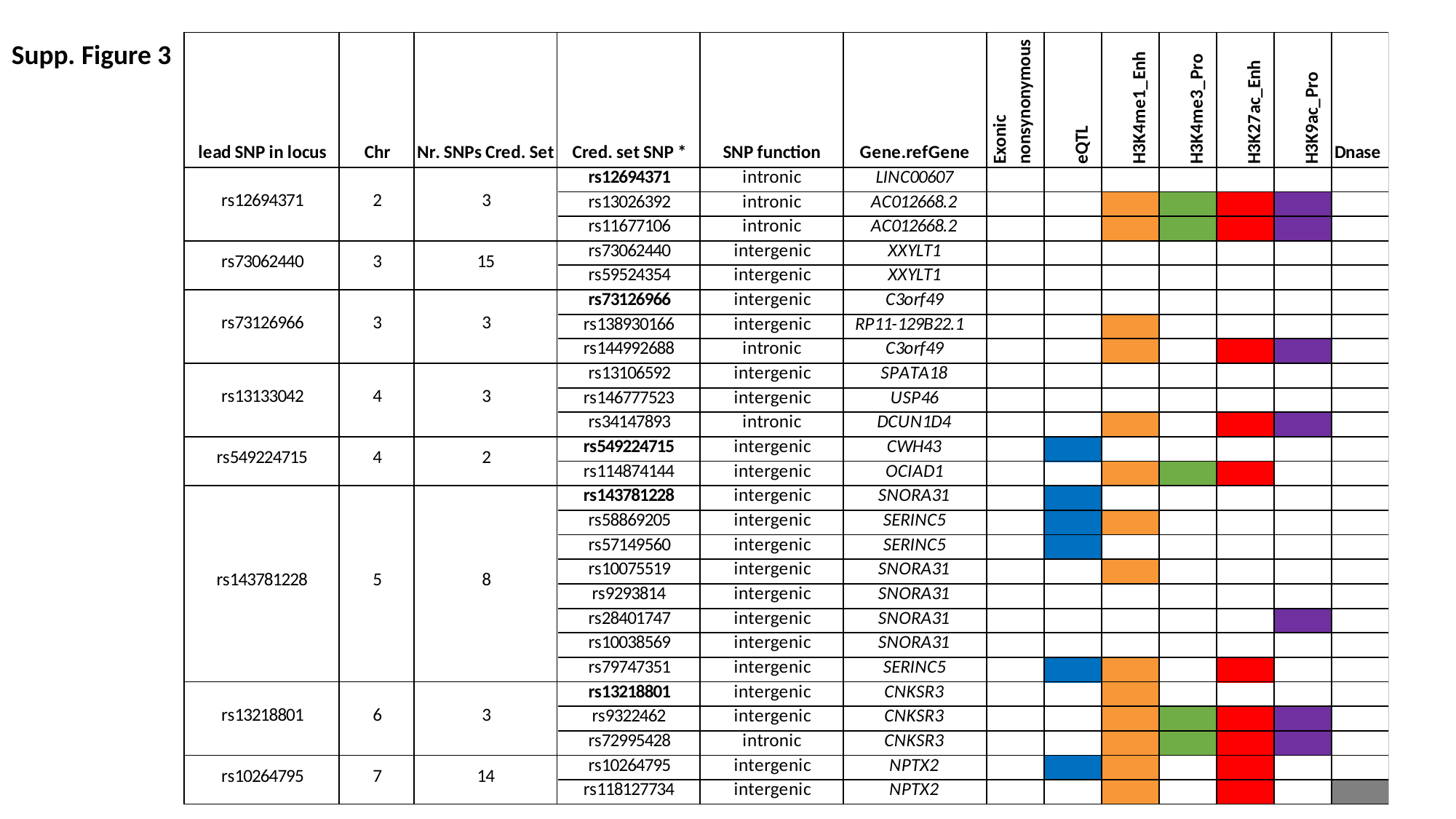

Supp. Figure 3

### Slide 4
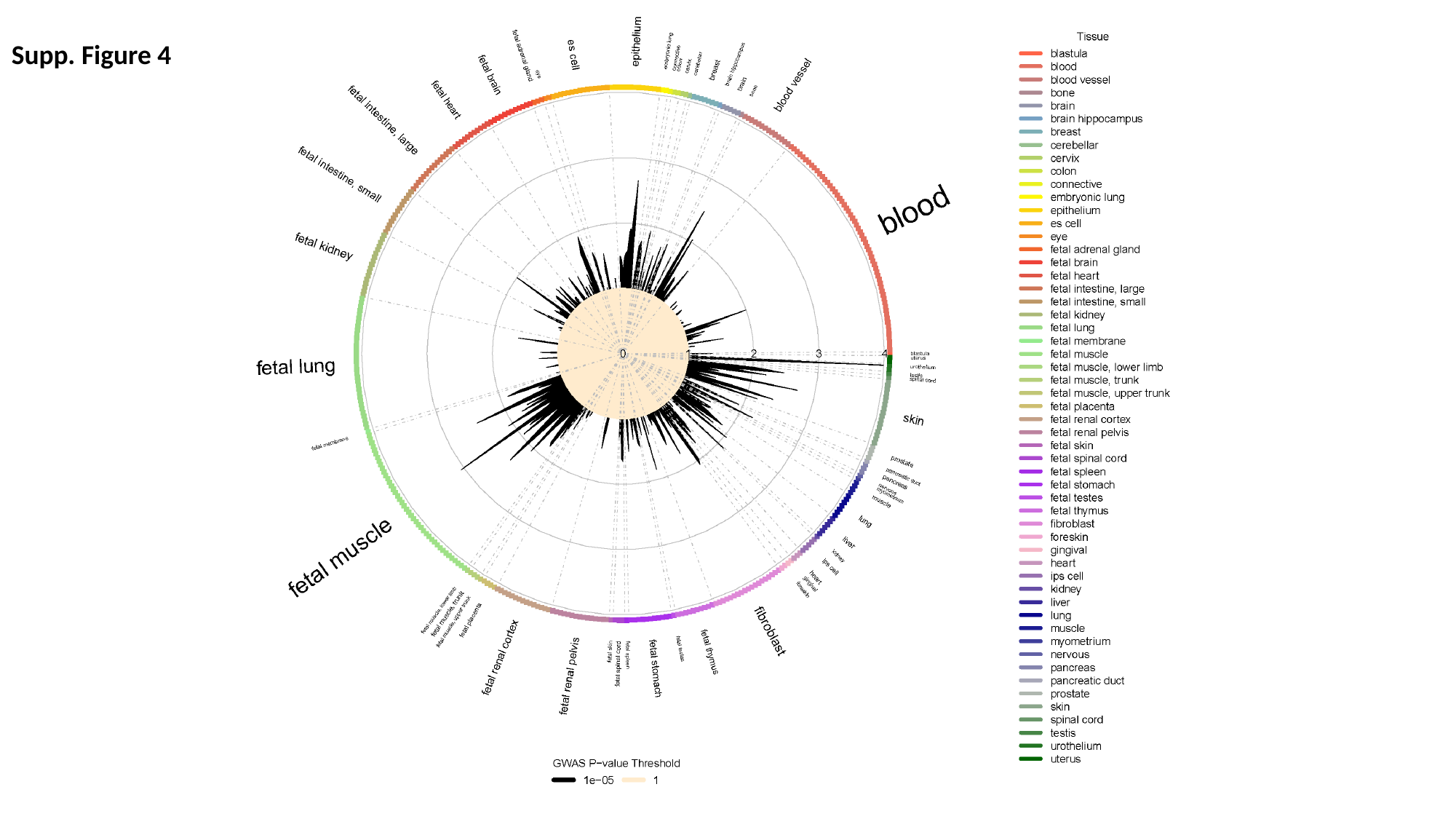

Supp. Figure 4

### Slide 5
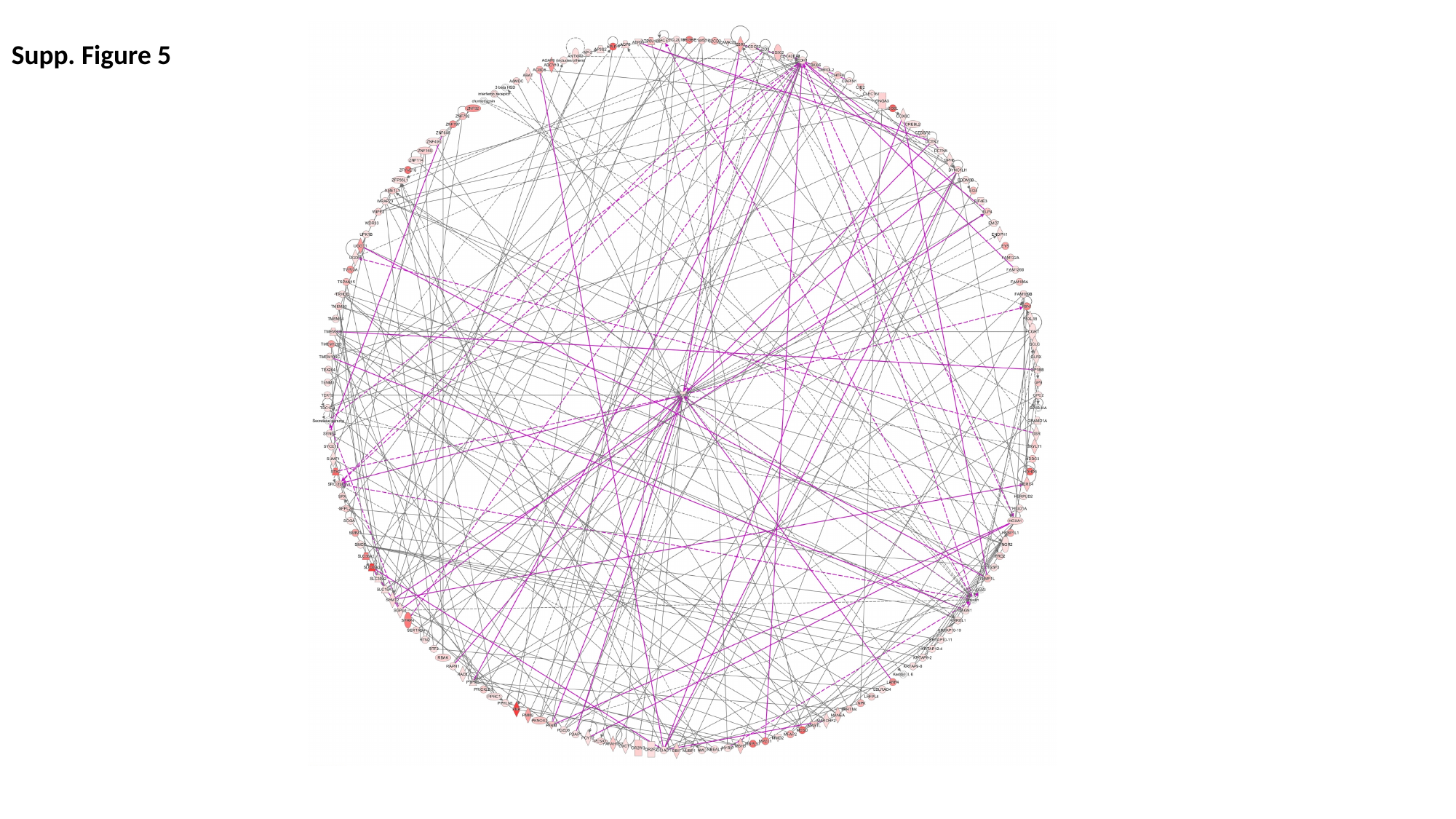

Supp. Figure 5
