## supplementary figure legend for "Genome-wide association study of nociceptive musculoskeletal pain treatment response in UK Biobank"

Supplementary Figure 1. Q-Q plot of A) pain treatment response for nociceptive musculoskeletal pain using ordinal phenotypes ; B) subtype analysis; C) secondary analysis.

Supplementary Figure 2. Manhattan plot of A) pain treatment response for nociceptive musculoskeletal pain using ordinal phenotypes; B) subtype analysis; C) secondary analysis.

Supplementary Figure 3. Functional annotation results of 95% credible set SNPs. Different colors indicate credible SNPs in LD with exonic nonsynonymous variants, overlap with active chromatin marks of active enhancer (H3K4me1, H3K27ac) or promoters (H3K4me3, H3K9ac), and sensitive to DNAse. The selected cell types for annotating overlap with chromatin marks and DNAse are provided in supplementary Table 1. eQTL overlap is obtained from FUMA. * When the 95% credible set included more than 10 likely causal variants, only lead SNP and SNP with maximum posterior probability (PP_max_) were selected for annotation. SNPs in bold indicate that the lead SNP remains maximum posterior probability.

Supplementary Figure 4. Results of the cell type and tissue enrichment using primary GWAS signals. Enrichment was conducted with variants passing the suggestive significance thresholds 1 × 10^-5^ in DNase I hypersensitive site ( DHS) hotspot regions of 424 cell types, which are sorted by tissues along the outside edge of the plot. The radial axis in black represents the fold enrichment (FE) for each cell types. Significant enrichment is indicated with dots along the inside edge of the plot. However, no significant enrichment was observed.

Supplementary Figure 5. Network analysis of the main GWAS genes with ingenuity pathway analysis (IPA). The proteins encoded by the 1457 genes with nominal significance (P-value < 0.05 ; the list of these genes are provided in Supplementary Table 2) were used for the network analysis. After merging the top 5 networks and generating a ‘radial’plot, EGFR is at the center of molecular interactions. All proteins in red/pink are encoded by genes present in the input dataset. The more red a protein is, the more significant the P-value for the input gene that encodes this protein is.
