## supplementary table and data legend for "Genome-wide association study of nociceptive musculoskeletal pain treatment response in UK Biobank"

Supplementary Table 1. Selected cells and tissues for SNP functional annotation in Haploreg.

Supplementary Table 2. P-value of genes calculated by MAGMA.

Supplementary Table 3. Significant associated pathways in pain treatment response from IPA. -log(p-value), negative logarithm of pathway enrichment P-value. Ratio, the ratio of genes from input file in this pathway. Molecules, genes from input file in this pathway.

Supplementary Table 4. Enriched networks in pain treatment response from IPA. Score, negative logarithm of network enrichment P-value. Focus molecules, genes with the most interactions to other genes which are connected together to form a network.

Supplementary Table 5. Genetic correlations with pain treatment response. rg, the estimated genetic correlation. se, the bootstrap standard error of the genetic correlation estimate. z, the z-statistic rg/se for testing whether the genetic correlation is significantly different from zero. p: p-value from the z-statistic.

Supplementary Table 6. Nominal significant genetic correlations enrichment. Nr. of insignificant genetic correlations (P > 0.05), number of tested traits with insignificant genetic correlations with pain treatment response; Nr. of nominal significant genetic correlations (P < 0.05), number of tested traits with nominal significant genetic correlations with pain treatment response.

Supplementary Table 7. Lead SNPs identified in subtype GWAS. CHR:POS physical position of the SNP, A1 effect allele, AF1 effect allele frequency, BETA (SE) effect size of SNP and standard error (SE).

Supplementary Table 8. Lead SNPs identified in secondary GWAS. CHR:POS physical position of the SNP, A1 effect allele, AF1 effect allele frequency, BETA (SE) effect size of SNP and standard error (SE).

Supplementary data 1. Selected nociceptive musculoskeletal diagnosis in GP diagnosis data. Abbreviations: read_3, read code version 3 of diagnosis; read_2, read code version 2 of diagnosis.

Supplementary data 2. Selected pain medications in GP prescriptions data. Abbreviations: read_2, read code version 2 for medications; dmd_code, dmd code for medications; BNF, British national formula (BNF) code for medications; ladder, corresponding WHO analgesic ladders.
